# Association of total and brain-derived Alzheimer’s disease plasma biomarkers with brain amyloid deposition in a community-based sample

**DOI:** 10.64898/2026.07.29.26359150

**Authors:** Muge Akinci, Froogh Aziz, Diana Guzman, Lina Cheung, Jian X. Kong, Stephanie Silver, Joseph Eimicke, Sabrina Simoes, Jeanne A. Teresi, Adam M. Brickman, Patrick Lao, José A. Luchsinger

**Affiliations:** Department of Medicine, College of Physicians and Surgeons, Columbia University Irving Medical Center, 630 West 168th Street, New York, NY 10032, United States; Taub Institute for Research on Alzheimer’s Disease and the Aging Brain, Vagelos College of Physicians and Surgeons, Columbia University, 710 West 168th Street, New York, NY 10032, United States; Morris Stroud III Center for Study of Quality of Life in Health and Aging, Columbia University, 1051 Riverside Drive, New York 10032, NY, United States; Gertrude H. Sergievsky Center, Vagelos College of Physicians and Surgeons, Columbia University, 630 West 168th Street, New York, NY 10032, United States; New York State Psychiatric Institute, 1051 Riverside Drive, New York 10032, NY, United States; Department of Neurology, Vagelos College of Physicians and Surgeons, Columbia University, 710 West 168th Street, New York, NY 10032, United States; Department of Epidemiology, Joseph P. Mailman School of Public Health, Columbia University Irving Medical Center, 722 West 168th Street, New York, NY 10032, United States

## Abstract

**Background and Objectives:** Plasma biomarkers, particularly brain-derived phosphorylated-tau (BD-p-tau) species, hold promise as screening tools for Alzheimer’s disease (AD). However, their ability to reflect AD pathology remains understudied in community settings. In a community-based sample, we examined associations between plasma biomarkers and cerebral amyloid (Aβ) deposition, and whether kidney function modified these associations.

**Methods:** This cohort study included cognitively unimpaired, late middle-aged adults with^18^F-Florbetaben PET imaging and NULISAseq-derived plasma biomarker measurements. Analyses were restricted to NULISAseq biomarkers related to AD pathology (Aβ38, Aβ40, Aβ42, ACHE, BACE1, BASP1, BD-p-tau181, BD-p-tau217, CD63, IGFBP7, KLK6, MAPT-tau, PSEN1, SFRP1, total p-tau181, p-tau217, and p-tau231). As a measure of kidney function, cystatin C–based estimated glomerular filtration rate was calculated and categorized by chronic kidney disease (CKD) stage as normal/high (≥90 mL/min/1.73 m^2^); mildly decreased (60–89 mL/min/1.73 m^2^); and moderately/severely decreased or failure (<60 mL/min/1.73 m^2^). Bidirectional stepwise linear regression analysis was performed to identify plasma biomarkers associated with brain Aβ deposition. Linear regression models including plasma biomarker-by-CKD stage interactions tested effect modification by kidney function.

**Results:** A total of 541 Hispanic, non-Hispanic Black, and non-Hispanic White participants were included. Stepwise linear regression retained plasma BD-p-tau217 (B = 0.22; 95% CI 0.19 to 0.25; *p* < 0.001), which was positively associated with brain Aβ deposition, alongside Aβ42 (B = −0.08; 95% CI −0.11 to −0.06; *p* < 0.001), IGFBP7 (B = −0.05; 95% CI −0.08 to −0.02; *p* = 0.004), and BACE1 (B = −0.04; 95% CI −0.07 to −0.01; *p* = 0.009), which were negatively associated with brain Aβ deposition. A significant BD-p-tau217-by-CKD stage interaction demonstrated a weaker association between BD-p-tau217 and brain Aβ deposition among individuals with moderately/severely decreased kidney function or failure than those with normal/high kidney function (B = −0.17; 95% CI −0.25 to −0.08; *p* < 0.001).

**Discussion:** In a real-world sample, BD-p-tau217 emerged as the plasma biomarker most strongly associated with brain Aβ deposition, although this association may be attenuated in the presence of moderate/severe kidney dysfunction or kidney failure. IGFBP7 and BACE1 were identified as candidate plasma biomarkers of brain Aβ deposition, warranting replication in independent cohorts.

## Introduction

Blood-based proteomic biomarkers have emerged as cost-effective tools for supporting risk assessment, early diagnosis, staging, and monitoring of Alzheimer’s disease (AD).^1,2^ The most promising plasma biomarkers for AD include amyloid-beta (Aβ)42, the Aβ42/40 ratio, and phosphorylated tau (p-tau) species—specifically p-tau181, p-tau217, and p-tau231.^3,4^ These biomarkers correlate well with positron emission tomography (PET) or cerebrospinal fluid (CSF) measures of Aβ pathology,^3–6^ the main feature of AD pathogenesis.^4^ Notably, p-tau217 is gaining recognition as the best-performing plasma biomarker for identifying AD pathology,^7,8^ demonstrating superior diagnostic accuracy compared with other plasma biomarkers.^9,10^ Indeed, plasma p-tau217 levels can increase very early in the course of AD,^11^ even in the presence of subtle cortical Aβ aggregation.^12,13^ Moreover, evidence suggests that plasma p-tau217—alone or in combination with the plasma Aβ42/40 ratio—can predict future Aβ pathology in cognitively unimpaired (CU) individuals with low baseline brain Aβ levels.^14^ Hence, plasma biomarkers could be useful for identifying individuals at risk of Aβ accumulation who may benefit from primary prevention strategies.^12,14^

Despite their high potential as large-scale screening tools for Aβ pathology,^14–17^ relatively few studies examined the associations between plasma biomarkers and brain Aβ deposition within the community at large.^18–20^ This is particularly relevant as the performance of plasma biomarkers in detecting early Aβ pathology may differ between clinic-based and community-based populations.^13^ Furthermore, compared with clinic-based cohorts, community-based cohorts tend to have a higher prevalence of medical comorbidities,^18,21^ which may also vary in prevalence across racial and ethnic groups.^22,23^ These comorbidities may alter plasma biomarker levels through peripheral physiological factors and thereby affect their interpretation.^23,24^ In particular, chronic kidney disease (CKD) has been consistently linked to elevated plasma biomarker concentrations,^23–26^ potentially influencing the total blood pool of these biomarkers through reduced glomerular filtration.^27^ Notably, most immunoassays for p-tau measure “total p-tau”^28,29^ as they recognize N-terminal epitopes common to both brain-derived, low-molecular-weight tau and high-molecular-weight tau expressed by the peripheral nervous system and other organs.^30–32^ Consequently, these assays may have limited ability to distinguish brain-specific, AD-related pathological changes from peripheral physiological alterations,^28,33^ including those associated with CKD.

Recently, assays targeting brain-derived tau isoforms have been developed to mitigate interference from peripheral sources and enhance specificity for brain-derived pathology.^28,34,35^ Studies utilizing these assays reported that brain-derived p-tau (BD-p-tau)217 and BD-p-tau181 had superior performance than total p-tau isoforms in identifying abnormal brain Aβ levels,^32,33^ with BD-p-tau217 further exhibiting reduced interference from kidney dysfunction.^33^ Nevertheless, the association of BD-p-tau isoforms with brain Aβ burden, and the extent to which kidney function may modify these associations, remain unclear in community settings. A better understanding of these relationships in more heterogeneous, community-based samples will guide the appropriate use and interpretation of plasma biomarkers for AD risk screening at the population level.

In the current study, we used a NUcleic acid Linked Immuno-Sandwich assay (NULISAseq)^35^ to quantify plasma biomarkers of AD pathology, including BD-p-tau isoforms, in a community-based, racially and ethnically diverse sample of CU adults. In this real-world cohort, our primary aim was to identify NULISAseq biomarkers most strongly associated with cerebral Aβ pathology. Our secondary aim was to examine whether CKD modifies the association between NULISAseq biomarkers and cerebral Aβ pathology.

## Methods

### Standard protocol approvals, registrations, and patient consents

In this cross-sectional study, 550 individuals with Aβ PET imaging data and available frozen plasma samples were considered for inclusion. Participants were recruited from the community surrounding Columbia University Irving Medical Center (CUIMC) in New York City, NY, USA (see ^36^ for recruitment details). Inclusion criteria were: (i) age between 55 and 70 years and (ii) willing and able to undergo clinical and neuroimaging assessments. Exclusion criteria were: (i) self-reported diagnosis of dementia, (ii) cancer other than nonmelanoma skin cancer, and (iii) contraindications to PET imaging.

Demographic, clinical, laboratory and imaging data were collected between 2016 and 2022. Sex was self-reported by participants in response to the question: “Are you female or male?” All participants reported being either male or female and were classified as men or women, respectively. Race and ethnicity information was also collected through self-report via a structured questionnaire. Categories reported by participants included Hispanic, non-Hispanic Black, and non-Hispanic White. *APOE* ε4 genotype was determined based on the single-nucleotide variants rs429358 and rs7412 (LGC Genomics). Participants carrying at least one ε4 allele were classified as *APOE* ε4 carriers and others as non-carriers.

Study protocols were approved by the Institutional Review Board and the Joint Radiation Safety Commission at Columbia University. Written informed consent was obtained from all participants.

### Measurement of kidney function

As a measure of kidney function, estimated glomerular filtration rate (eGFR) was derived using cystatin C levels. Cystatin C was quantified in blood samples collected at CUIMC using a particle-enhanced turbidimetric assay on the automated analyzer Integra 400 plus (Roche Diagnostics, Indianapolis, IN, USA). The assay had a lower limit of detection of 0.4 mg/L. The intra- and inter-assay precision values were 1.58% and 2%, respectively. eGFR was calculated using the 2012 Chronic Kidney Disease Epidemiology Collaboration (CKD-EPI) cystatin C equation,^37^ based on plasma cystatin C levels, age, and sex, as follows: 133 × min(Scys/0.8, 1)^−0.499^ × max(Scys/0.8, 1)^−1.328^ × 0.996^Age^ × 0.932 (if female).

eGFR was evaluated both as a continuous measure and as a categorical variable based on CKD stage. According to the Kidney Disease: Improving Global Outcomes Chronic Kidney Disease Guideline Development Work Group criteria,^38^ CKD is classified into five stages based on eGFR: (i) eGFR ≥90 mL/min/1.73 m^2^ indicating possible kidney damage with normal or high kidney function; (ii) 60–89 mL/min/1.73 m^2^ indicating kidney damage with mild loss of kidney function; (iii) 30–59 mL/min/1.73 m^2^ indicating moderate loss of kidney function; (iv) 15–29 mL/min/1.73 m^2^ indicating severe loss of kidney function; and (v) <15 mL/min/1.73 m^2^ indicating kidney failure. Due to sparse data in the moderate/severe loss of kidney function and kidney failure stages, a three-category CKD stage variable reflecting kidney function status was created: (i) normal/high kidney function (eGFR ≥90 mL/min/1.73 m^2^); (ii) mildly decreased kidney function (eGFR 60–89 mL/min/1.73 m^2^); and (iii) moderately/severely decreased kidney function or failure (eGFR <60 mL/min/1.73 m^2^).

### NULISAseq analysis and data processing

Plasma samples were collected at CUIMC and sent to Alamar Biosciences, Inc (Fremont, CA, USA) for NULISAseq measurements. Samples were stored at −80 °C until ready for use. Before the assay, samples were thawed and centrifuged at 2,200g for 10 min. 25 μL supernatant from the sample was plated and analyzed with the NULISAseq™ 120 CNS disease panel. Samples were incubated with a paired set of capture and detection antibodies. Capture antibodies were conjugated with partially double-stranded DNA which contains a poly-A tail and a target-specific molecular identifier. Detection antibodies were conjugated with another partially double-stranded DNA that contains a biotin group and a matching target-specific barcode. Incubation of the antibodies resulted in the formation of immunocomplexes, which were then captured by addition of paramagnetic oligo-dT beads with dT-polyA hybridization, washed, released, recaptured, and underwent a second round of wash to identify intact immunocomplexes. DNA reporter molecules containing unique target-specific molecular identifiers (TMI) and sample-specific molecular identifiers (SMI) were generated through ligation and quantified by Next-Generation Sequencing (NGS).^35^

Assay controls were run (i.e., sample control, internal control, inter-plate control, and negative control) and data normalization was performed. In brief, internal control normalization was performed by dividing each sample and target read count by that sample’s internal control counts. Then, inter-plate control normalization was performed by dividing internal control-normalized counts by target-specific medians of inter-plate controls. The data were added by 1 and log2-transformed to form a more normal distribution. These values are referred to as NULISA Protein Quantification (NPQ) units. Plate-specific limit of detection (LOD) was calculated for target assays by taking the mean plus three times the standard deviation of the unlogged normalized counts for the negative control samples on the plate, which were then rescaled and log2-transformed. A target was considered detectable if the target detectability (i.e., the percentage of samples above the LOD) exceeded > 50%.

For statistical analyses, NULISAseq proteins linked to Aβ and tau pathologies were considered as independent variables. These included Aβ peptides (Aβ38, Aβ40, and Aβ42), acetylcholinesterase (ACHE), beta-secretase 1 (BACE1), brain abundant membrane attached signal protein 1 (BASP1), CD63 molecule (CD63), insulin-like growth-factor binding protein 7 (IGFBP7), kallikrein-6 (KLK6), presenilin-1 (PSEN1), secreted frizzled-related protein 1 (SFRP1), total p-tau isoforms (p-tau181, p-tau217, and p-tau231), BD-p-tau isoforms (BD-p-tau181 and BD-p-tau217), and total microtubule-associated protein tau (MAPT-tau). Despite being included in the NULISA panel for Aβ and tau pathologies, four proteins were excluded from the analyses: BD-p-tau231 and BD-MAPT-tau due to low detectability (<50%); cystatin C (CST3), as it was used for eGFR calculation; and *APOE* protein, since APOE genotyping was used to assess APOE-related genetic risk.

### Aβ PET imaging acquisition and processing

Cerebral Aβ burden was quantified using ^18^F-Florbetaben PET acquired on a Siemens Biograph mCT 64 scanner. The target injected dose was 8.1 mCi (± 10%). Images were acquired at 90–110 minutes postinjection in 5-minute frames and were reconstructed using an iterative reconstruction algorithm with a voxel size of 1.6 × 1.6 × 2.0 mm^3^. The standardized uptake value ratio (SUVR) was calculated in native space using the inferior cerebellar gray matter as the reference region.^39^

As the outcome of interest, a global Aβ composite was derived by averaging FreeSurfer-defined composite Thal phase^40^ regions, encompassing the frontal cortex, temporal cortex, parietal cortex, cingulate cortex, and striatum. Aβ positivity was defined using a global Aβ SUVR cutoff of 1.25 (≥1.25, Aβ-positive; <1.25, Aβ-negative).^39^

### Statistical analysis

Differences across CKD stages were evaluated using the Kruskal–Wallis H test for continuous variables and Fisher’s exact test for categorical variables. Since *APOE* ε4 is a predictor of Aβ pathology,^41^ plasma biomarker levels were compared between *APOE* ε4 carriers and non-carriers using the Wilcoxon rank sum test. Spearman correlations were performed to examine (i) intercorrelations among individual NULISAseq plasma biomarkers; (ii) correlations between each plasma biomarker and eGFR; and (iii) correlations between each plasma biomarker and global Aβ SUVR. For statistically significant correlations, partial Spearman analyses adjusted for eGFR were performed to assess whether the correlations between plasma biomarkers and global Aβ SUVR remained independent of kidney function.

In the main analysis, bidirectional stepwise linear regression was performed to identify a parsimonious set of plasma biomarkers associated with global Aβ SUVR. Prior to model selection, multicollinearity among the biomarkers was assessed using the variance inflation factor (VIF), and biomarkers with high collinearity (VIF ≥ 5) were excluded from the stepwise procedure. The criteria for entry and removal in the model were set at p ≤ 0.05 and p ≥ 0.10, respectively. Goodness of fit was evaluated using adjusted *R^2^* and the partial F-test. Linear regression analyses including interaction terms between the identified plasma biomarkers and CKD stages were performed to assess effect modification by kidney function status.

As an exploratory analysis, stepwise logistic regression was performed to identify plasma biomarkers associated with Aβ PET positivity. Model fit was assessed using Nagelkerke *R^2^* and the likelihood ratio test. The receiver operating characteristic (ROC) analysis was conducted to evaluate the performance of selected biomarkers for detecting Aβ positivity. Area under the curve (AUC) values were calculated and compared using the DeLong test.

In sensitivity analyses, stepwise linear regression was performed with adjustment for age, as older age is a risk factor for both AD^41^ and CKD.^38^ Additionally, the least absolute shrinkage and selection operator (LASSO) method was applied as an alternative variable selection approach to bidirectional stepwise regression. To determine the optimal tuning parameter (λ), a 10-fold external cross-validation scheme was implemented using a fixed random seed for reproducibility.

Statistical analyses were conducted using R version 4.4.3 (R Foundation for Statistical Computing) and SAS 9.4 software. A two-sided *p* < .05 was considered statistically significant.

### Data availability

Anonymized data not published within this article will be made available by request from any qualified investigator.

## Results

Of the 550 participants with Aβ PET scans and frozen plasma, 4 (0.4%) had unusable Aβ PET data, and 5 (0.5%) did not pass the quality check for NULISAseq panel. Therefore, the final sample comprised 541 participants with available Aβ PET and plasma biomarkers data.

Participant characteristics for the entire sample and by CKD stage are summarized in **Table 1**. One hundred seventy-four (32.1%) participants had normal/high kidney function, 286 (52.9%) had mildly decreased kidney function, and 81 (15.0%) had moderately/severely decreased kidney function or failure, of whom 77 (95%) had moderate loss of kidney function, 2 (2.5%) had severe loss of kidney function, and 2 (2.5%) had kidney failure. The three CKD groups had significant differences in age (Kruskal–Wallis H statistic, H[2] = 33.66; *p <* .001), cystatin C (H[2] = 428.87; *p <* .001), plasma Aβ38 (H[2] = 32.07; *p <* .001), Aβ40 (H[2] = 87.97; *p <* .001), Aβ42 (H[2] = 95.70; *p <* .001), BD-p-tau181 (H[2] = 47.86; *p <* .001), BD-p-tau217 (H[2] = 18.06; *p <* .001), IGFBP7 (H[2] = 52.04; *p <* .001), KLK6 (H[2] = 47.82; *p <* .001), MAPT-tau (H[2] = 54.86; *p <* .001), total p-tau181 (H[2] = 41.19; *p <* .001), total p-tau217 (H[2] = 54.86; *p <* .001), and total p-tau231 (H[2] = 52.44; *p <* .001), with values highest among participants with moderately/severely decreased kidney function or failure, followed by those with mildly decreased kidney function and those with normal/high kidney function. In contrast, eGFR was highest among individuals with normal/high kidney function, followed by those with mildly decreased kidney function and those with moderately/severely decreased kidney function or failure (H[2] = 440.44; *p <* .001). There were no statistically significant group differences in years of education, sex, race and ethnicity, *APOE* ε4 status, global Aβ SUVR, Aβ PET positivity, and plasma ACHE, BACE1, BASP1, CD63, and SFRP1 levels (**Table 1**). Compared to *APOE* ε4 non-carriers, *APOE* ε4 carriers had significantly higher plasma total p-tau181 (Wilcoxon statistic, W = 29; *p* = .014), total p-tau217 (W = 26; *p* < .001), total p-tau231 (W = 28; *p* < .01), BD-p-tau181 (W = 27; *p* < .001) and BD-p-tau217 (W = 25; *p* < .001) levels, as well as lower plasma Aβ42 (W = 26; *p* < .001) levels.

**Table 1.** Participant characteristics in the entire sample and stratified by normal/high (eGFR≥90), mildly decreased (eGFR 60–89) and moderately/severely decreased kidney function or failure (eGFR<60) categories.

| Variable | Entire<br>Sample<br>N = 541 | Normal/high<br>kidney function<br>n = 174 | Mildly decreased<br>kidney function<br>n = 286 | Moderately/severely decreased<br>kidney function or failure<br>n = 81 | p value |
| --- | --- | --- | --- | --- | --- |
| Age, mean ± SD, years | 64.20 ± 3.18 | 63.15 ± 2.89 | 64.51 ± 3.14 | 65.42 ± 3.30 | <.001 |
| Education, mean ± SD, years | 11.99 ± 4.08 | 12.57 ± 3.74 | 11.76 ± 4.27 | 11.52 ± 4.03 | .053 |
| Women, no. (%) | 351 (64.9) | 113 (64.9) | 190 (66.4) | 48 (59.3) | .473 |
| Hispanic, no. (%) | 344 (63.6) | 113 (64.9) | 184 (64.3) | 47 (58.0) | .354 |
| Non-Hispanic Black, no. (%) | 120 (22.2) | 33 (19.0) | 68 (23.8) | 19 (23.5) |  |
| Non-Hispanic White, no. (%) | 77 (14.2) | 28 (16.1) | 34 (11.9) | 15 (18.5) |  |
| APOE-ε4 carrier, no. (%) | 189 (34.9) | 53 (30.5) | 109 (38.1) | 27 (33.3) | .243 |
| Global Aβ SUVR, mean ± SD | 1.17 ± 0.12 | 1.16 ± 0.12 | 1.18 ± 0.13 | 1.16 ± 0.10 | .107 |
| Aβ PET positive, no. (%) | 85 (15.7) | 23 (13.2) | 50 (17.5) | 12 (14.8) | .474 |
| Plasma Cystatin C, mean ± SD | 1.08 ± 0.43 | 0.79 ± 0.06 | 0.99 ± 0.08 | 1.49 ± 0.95 | <.001 |
| eGFR, mean ± SD | 78.70 ± 17.80 | 98.15 ± 5.79 | 75.08 ± 7.95 | 49.70 ± 10.89 | <.001 |
| Aβ38, NPQ, mean ± SD | 10.30 ± 0.89 | 10.08 ± 0.87 | 10.33 ± 0.86 | 10.68 ± 0.87 | <.001 |
| Aβ40, NPQ, mean ± SD | 11.29 ± 0.70 | 11.01 ± 0.59 | 11.30 ± 0.66 | 11.81 ± 0.74 | <.001 |
| Aβ42, NPQ, mean ± SD | 13.08 ± 0.42 | 12.90 ± 0.34 | 13.07 ± 0.36 | 13.47 ± 0.50 | <.001 |
| ACHE, NPQ, mean ± SD | 12.67 ± 0.42 | 12.67 ± 0.42 | 12.66 ± 0.42 | 12.68 ± 0.40 | .538 |
| BACE1, NPQ, mean ± SD | 13.29 ± 0.29 | 13.31 ± 0.33 | 13.29 ± 0.27 | 13.25 ± 0.28 | .356 |
| BASP1, NPQ, mean ± SD | 12.10 ± 1.85 | 12.17 ± 1.90 | 12.06 ± 1.81 | 12.09 ± 1.88 | .557 |
| BD-p-tau181, NPQ, mean ± SD | 13.08 ± 0.56 | 12.83 ± 0.43 | 13.05 ± 0.47 | 13.38 ± 0.84 | <.001 |
| BD-p-tau217, NPQ, mean ± SD | 11.51 ± 0.32 | 11.44 ± 0.21 | 11.52 ± 0.28 | 11.64 ± 0.52 | <.001 |
| CD63, NPQ, mean ± SD | 12.41 ± 0.56 | 12.36 ± 0.57 | 12.40 ± 0.51 | 12.57 ± 0.69 | .092 |
| IGFBP7, NPQ, mean ± SD | 12.95 ± 0.33 | 12.85 ± 0.27 | 12.94 ± 0.31 | 13.19 ± 0.40 | <.001 |
| KLK6, NPQ, mean ± SD | 12.11 ± 0.44 | 11.97 ± 0.39 | 12.11 ± 0.37 | 12.43 ± 0.60 | <.001 |
| Total MAPT-tau, NPQ, mean ± SD | 11.89 ± 0.55 | 11.67 ± 0.49 | 11.91 ± 0.45 | 12.25 ± 0.77 | <.001 |
| PSEN1, NPQ, mean ± SD | 11.65 ± 0.51 | 11.47 ± 0.46 | 11.63 ± 0.43 | 12.08 ± 0.62 | <.001 |
| Total p-tau181, NPQ, mean ± SD | 12.53 ± 0.63 | 12.34 ± 0.61 | 12.56 ± 0.55 | 12.86 ± 0.79 | <.001 |
| Total p-tau217, NPQ, mean ± SD | 10.69 ± 0.54 | 10.50 ± 0.45 | 10.73 ± 0.45 | 11.00 ± 0.78 | <.001 |
| Total p-tau231, NPQ, mean ± SD | 12.64 ± 0.59 | 12.42 ± 0.50 | 12.67 ± 0.51 | 12.98 ± 0.80 | <.001 |
| SFRP1, NPQ, mean ± SD | 14.51 ± 1.19 | 14.49 ± 1.05 | 14.54 ± 1.27 | 14.44 ± 1.20 | .922 |

Data are presented as mean ± standard deviation for continuous variables and as number (percentage) for categorical variables. Abbreviations: Aβ , amyloid-beta; PET, positron emission tomography; SUVR, standardized uptake value ratio; eGFR, estimated glomerular filtration rate; NPQ, NULISA protein quantification, ACHE, acetylcholinesterase; BACE1, beta-secretase 1; BASP1, brain abundant membrane attached signal protein 1; CD63, CD63 molecule; IGFBP7, insulin-like growth-factor binding protein 7, KLK6, kallikrein-6; PSEN1, presenilin-1; SFRP1, secreted frizzled-related protein 1; p-tau, phosphorylated tau; BD-p-tau, brain-derived p-tau; MAPT-tau, microtubule-associated protein tau.

Regarding intercorrelations among plasma biomarkers, BASP1 did not evidence significant correlations with any other biomarker. Similarly, correlations between Aβ42 and SFRP1, and between BACE1 and PSEN1, were not significant. All other plasma biomarkers were positively correlated with one another. Except for ACHE, BACE, BASP1, and SFRP1, all plasma biomarkers were inversely correlated with eGFR (**Figure 1**). Further, BD-p-tau217, BD-p-tau181, total p-tau217, Aβ42, and IGFBP7 were significantly correlated with global Aβ SUVR (**Figure 2**). These correlations remained significant in partial correlation analyses adjusted for eGFR, with BD-p-tau217 (ρ = 0.22, *p* < .001), BD-p-tau181 (ρ = 0.16, *p* < .001), and total p-tau217 (ρ = 0.12, *p* < .001) evidencing positive correlations, and Aβ42 (ρ = −0.19, *p* < .001) and IGFBP7 (ρ = −0.10, *p* = .015) evidencing negative correlations with global Aβ SUVR.

**Figure 1.**
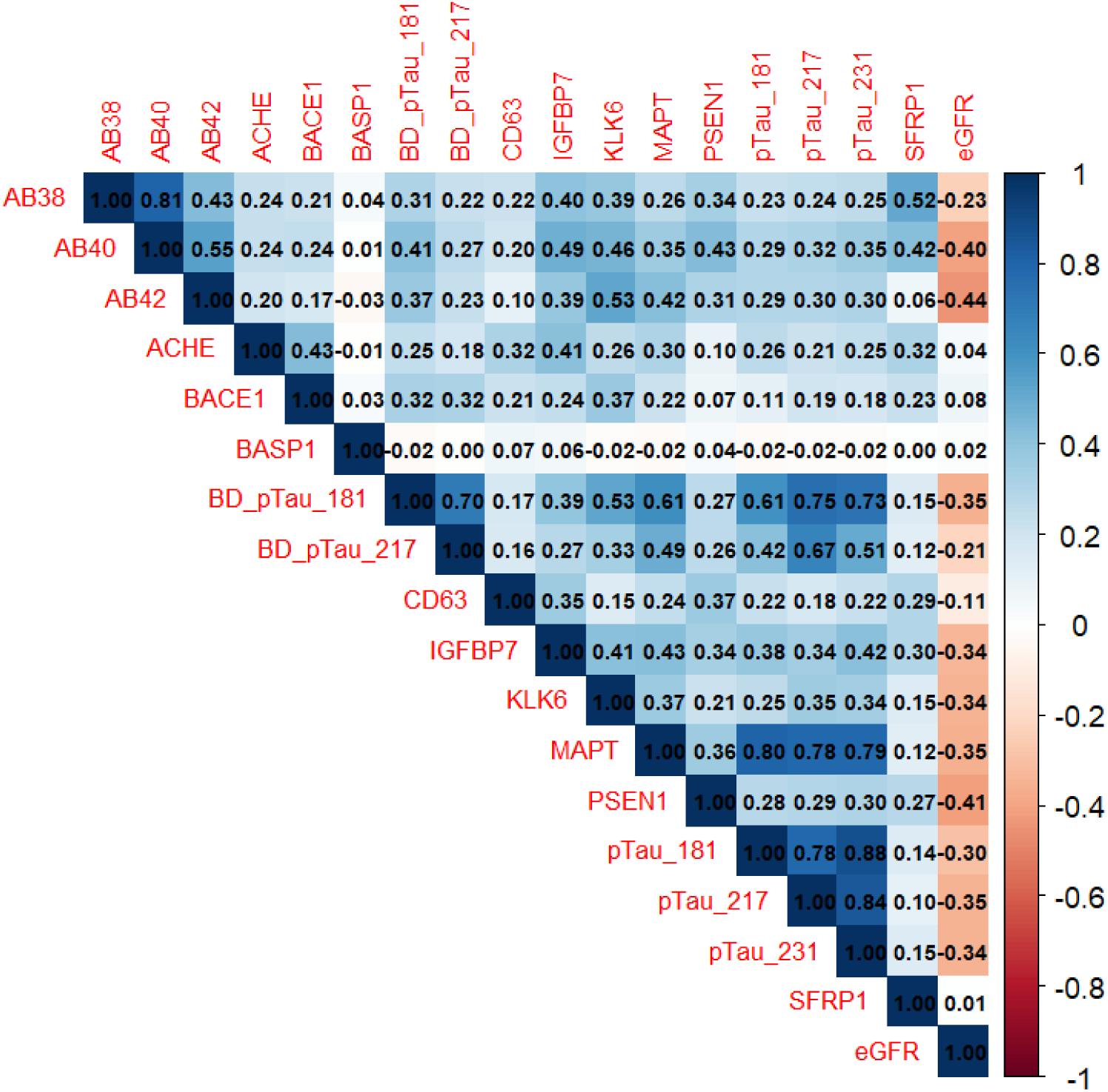
Spearman correlation matrix of NULISAseq-derived plasma biomarkers and eGFR. The heatmap illustrates pairwise Spearman correlation coefficients between plasma biomarkers. Color gradients indicate the magnitude and direction of the correlations (blue = positive correlation; red = negative correlation; range = −1 to 1), with darker shades representing stronger correlations. Abbreviations: Aβ, amyloid-beta; SUVR, standardized uptake value ratio; ACHE, acetylcholinesterase; BACE1, beta-secretase 1; BASP1, brain abundant membrane attached signal protein 1; CD63, CD63 molecule; IGFBP7, insulin-like growth-factor binding protein 7; KLK6, kallikrein-6; PSEN1, presenilin-1; SFRP1, secreted frizzled-related protein 1; p-tau, phosphorylated tau; BD-p-tau, brain-derived p-tau; MAPT-tau, microtubule-associated protein tau; eGFR, estimated glomerular filtration rate.

**Figure 2.**
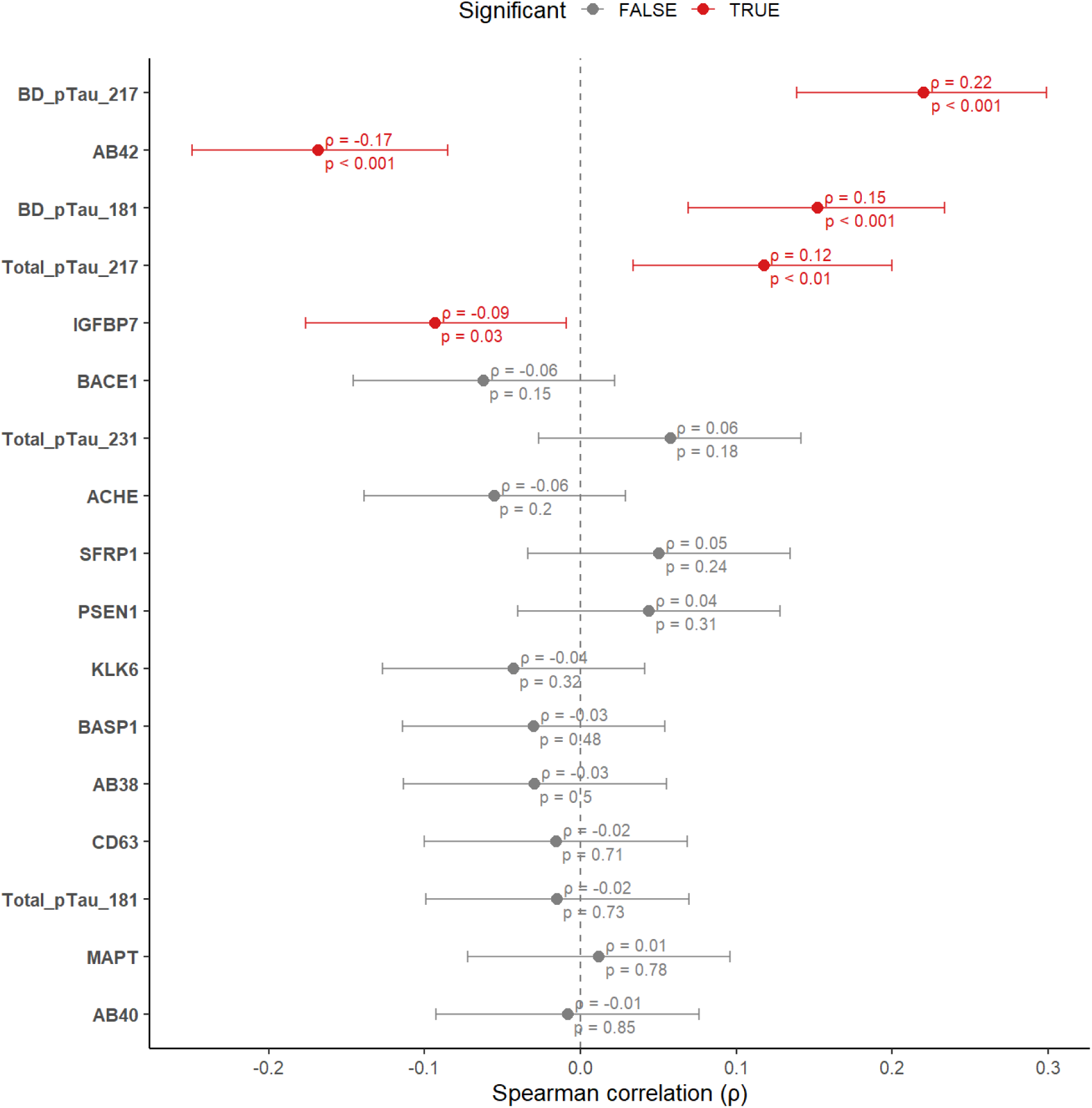
Forest plot showing the results from Spearman correlation analyses between plasma biomarkers and global Aβ SUVR. Spearman correlation coefficients between plasma biomarkers and global amyloid PET SUVR are presented with 95% confidence intervals and p values. Red points indicate statistically significant correlations (p < 0.05), while gray points indicate non-significant correlations. Abbreviations: Aβ, amyloid-beta; SUVR, standardized uptake value ratio; ACHE, acetylcholinesterase; BACE1, beta-secretase 1; BASP1, brain abundant membrane attached signal protein 1; CD63, CD63 molecule; IGFBP7, insulin-like growth-factor binding protein 7; KLK6, kallikrein-6; PSEN1, presenilin-1; SFRP1, secreted frizzled-related protein 1; p-tau, phosphorylated tau; BD-p-tau, brain-derived p-tau; MAPT-tau, microtubule-associated protein tau.

### Stepwise-selected plasma biomarkers associated with brain Aβ burden

Evaluation of multicollinearity among the plasma biomarkers revealed VIF values ≥ 5 for several tau biomarkers including total p-tau181 (VIF = 6.18), total p-tau217 (VIF = 8.21), total p-tau231 (VIF = 8.10), and BD-p-tau181 (VIF = 7.30), which were excluded from the stepwise selection procedure. In contrast, BD-p-tau217 had a VIF value of 4.53 and was therefore retained in the model. Among the 13 remaining NULISAseq biomarkers, 4 were retained in the final stepwise model. Plasma BD-p-tau217 was selected first and demonstrated the strongest association, followed by Aβ42, IGFBP7, and BACE1. Higher levels of BD-p-tau217, and lower levels of Aβ42, IGFBP7, and BACE1 were associated with greater global Aβ SUVR (**Table 2**). Compared with the model including BD-p-tau217 alone, the addition of Aβ42, IGFBP7, and BACE1 significantly improved model fit (adjusted *R*^2^ change = 0.120; F[3, 536] = 31.42, *p* < .001).

**Table 2.** Stepwise selection of NULISAseq plasma biomarkers associated with global Aβ SUVR.

| **Outcome:** Global Aβ SUVR | | | |
| --- | --- | --- | --- |
| **Plasma Biomarker** | **B (95% CI)** | ***p* value** | **Adjusted *R*<sup>2</sup>** |
| BD-p-tau217 | 0.22 (0.19 to 0.25) | <.001 | .167 |
| Aβ42 | −0.08 (−0.11 to −0.06) | <.001 | .265 |
| IGFBP7 | −0.05 (−0.08 to −0.02) | .004 | .280 |
| BACE1 | −0.04 (−0.07 to −0.01) | .009 | .287 |

Plasma biomarkers selected by stepwise linear regression are listed in the order of selection. Adjusted *R*<sup>2</sup> values represent the performance of the cumulative model at each step. Abbreviations: Aβ , amyloid-beta; SUVR, standardized uptake value ratio; BD-p-tau, brain-derived p-tau; IGFBP7, insulin-like growth-factor binding protein 7; BACE1, beta-secretase 1.

### Interactions between plasma biomarkers and CKD stage on brain Aβ burden

Among the 4 plasma biomarkers associated with global Aβ SUVR, only BD-p-tau217 interacted with CKD stage on global Aβ SUVR (**Table 3**). Hence, we repeated the model including only the interaction term between BD-p-tau217 and CKD stage. Results showed that the association between BD-p-tau217 and global Aβ SUVR was significantly weaker in participants with moderately/severely decreased kidney function or failure than in those with normal/high kidney function (**Table 4 & Figure 3**). However, this observation was limited by the relatively small sample size of the group with moderately/severely decreased kidney function or failure (n = 81), compared to the groups with normal/high (n = 174) and mildly decreased (n = 286) kidney function. Thus, we conducted a post-hoc analysis and repeated the interaction analysis, grouping participants with any degree of kidney dysfunction (i.e., mildly, moderately, severely decreased kidney function or failure, n = 367) together and comparing them with those who had normal/high kidney function (n = 174). The interaction term was not significantly associated with global Aβ SUVR (**Supplementary Table 1**).

**Figure 3.**
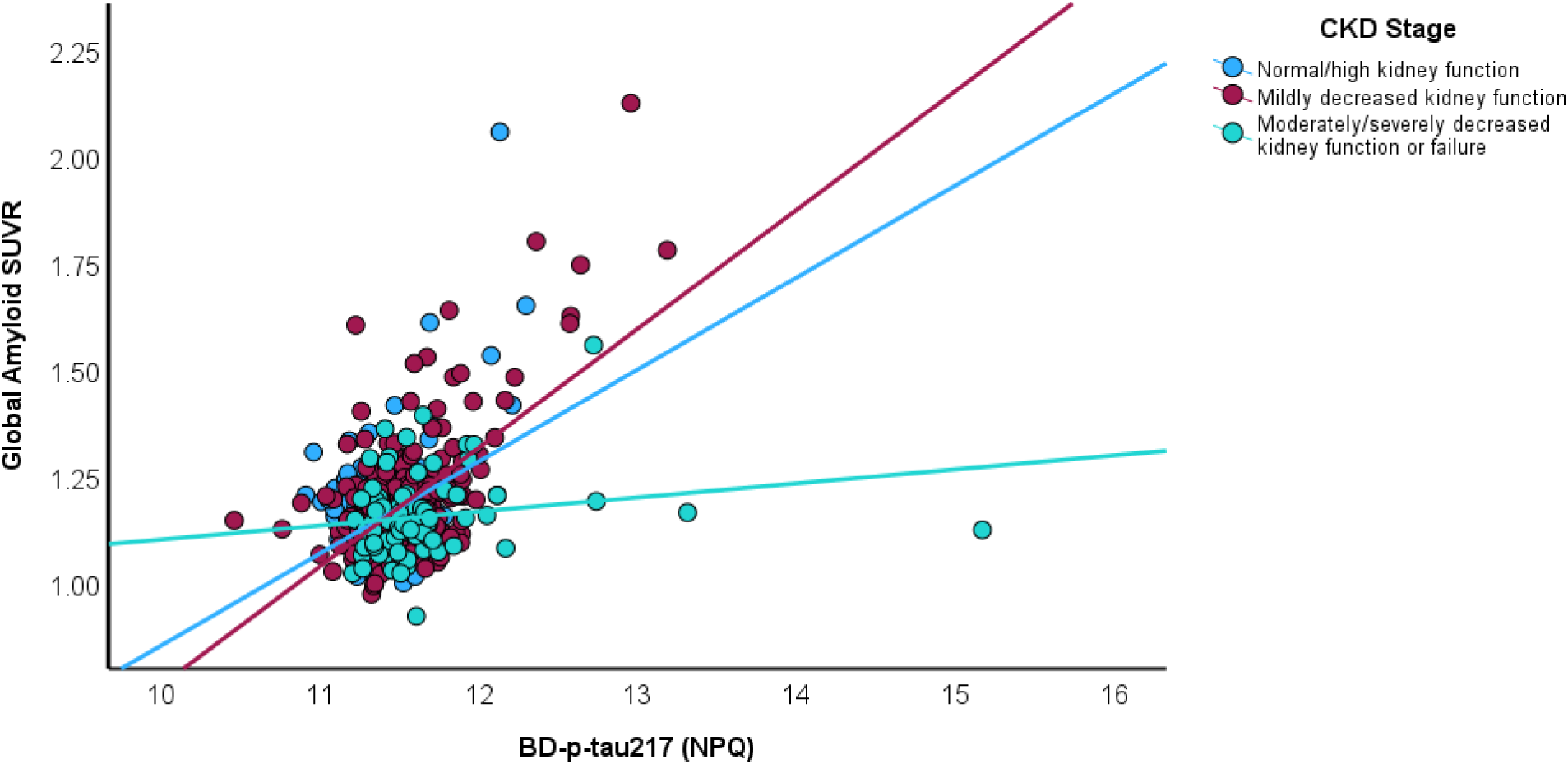
Interaction between plasma BD-p-tau217 and CKD stage on brain Aβ burden. The figure illustrates the relationship between BD-p-tau217 and brain amyloid-β burden by kidney function status. Abbreviations: SUVR, standardized uptake value ratio; NPQ, NULISA protein quantification; BD-p-tau, brain-derived p-tau.

**Table 3.** Linear regression results for interactions between stepwise-selected plasma biomarkers and CKD stage on global Aβ SUVR.

| Independent Variable | Outcome: Global Aβ SUVR | |
| --- | --- | --- |
|  | B (95% CI) | p value |
| BD-p-tau217*CKD stage (mildly decreased kidney function) | 0.05 (−0.04 to 0.13) | .316 |
| BD-p-tau217*CKD stage (moderately/severely decreased kidney function or failure) | −0.16 (−0.25 to −0.06) | .002 |
| Aβ42*CKD stage (mildly decreased kidney function) | 0.05 (−0.006 to 0.11) | .078 |
| Aβ42*CKD stage (moderately/severely decreased kidney function or failure) | 0.05 (−0.03 to 0.13) | .218 |
| IGFBP7*CKD stage (mildly decreased kidney function) | −0.01 (−0.09 to 0.06) | .783 |
| IGFBP7*CKD stage (moderately/severely decreased kidney function or failure) | −0.01 (−0.11 to 0.09) | .790 |
| BACE1*CKD stage (mildly decreased kidney function) | −0.03 (−0.10 to 0.04) | .348 |
| BACE1*CKD stage (moderately/severely decreased kidney function or failure) | −0.04 (−0.14 to 0.06) | .400 |
| CKD stage (mildly decreased kidney function) | 0.02 (−0.002 to 0.04) | .080 |
| CKD stage (moderately/severely decreased kidney function or failure) | 0.01 (−0.02 to 0.05) | .443 |
| BD-p-tau217 | 0.26 (0.18 to 0.34) | <.001 |
| Aβ42 | −0.11 (−0.16 to −0.06) | <.001 |
| IGFBP7 | −0.03 (−0.09 to 0.04) | .421 |
| BACE1 | −0.02 (−0.08 to 0.03) | .345 |

Abbreviations: Aβ , amyloid-beta; CKD, chronic kidney disease; BD-p-tau, brain-derived p-tau; IGFBP7, insulin-like growth-factor binding protein 7; BACE1, beta-secretase 1.

**Table 4.** Linear regression results for the interaction between plasma BD-p-tau217 and CKD stage on global Aβ SUVR.

| Independent variable | Outcome: Global Aβ SUVR | |
| --- | --- | --- |
|  | B (95% CI) | p value |
| BD-p-tau217*CKD stage (mildly decreased kidney function) | 0.03 (−0.05 to 0.11) | .410 |
| BD-p-tau217*CKD stage (moderately/severely decreased kidney function or failure) | −0.17 (−0.25 to −0.08) | <.001 |
| CKD stage (mildly decreased kidney function) | 0.01 (−0.01 to 0.03) | .227 |
| CKD stage (moderately/severely decreased kidney function or failure) | 0.01 (−0.02 to 0.04) | .500 |
| BD-p-tau217 | 0.27 (0.19 to 0.34) | <.001 |
| Aβ42 | −0.07 (−0.10 to −0.05) | <.001 |
| IGFBP7 | −0.03 (−0.07 to −0.004) | .028 |
| BACE1 | −0.05 (−0.08 to −0.02) | .002 |

Abbreviations: Aβ , amyloid-beta; CKD, chronic kidney disease; BD-p-tau, brain-derived p-tau; IGFBP7, insulin-like growth-factor binding protein 7; BACE1, beta-secretase 1.

### Exploratory analysis: Stepwise-selected plasma biomarkers associated with Aβ PET positivity

Of the 13 plasma biomarkers evaluated, BD-p-tau217, Aβ42, and BACE1 were retained in the final stepwise logistic regression model. Higher plasma BD-p-tau217 levels, and lower plasma Aβ42 and BACE1 levels were associated with higher odds of being Aβ PET positive (**Supplementary Table 2)**. Compared with the model including plasma BD-p-tau217 alone, the addition of plasma Aβ42 and BACE1 significantly improved model fit (Nagelkerke *R*^2^ change = 0.112; χ^2^[3] = 71.46, *p* < .001). Results from the ROC analysis showed that the AUC value for BD-p-tau217 alone was 0.68 (95 CI%, 0.61 to 0.76), whereas the combined model yielded an AUC of 0.75 (95 CI%, 0.69 to 0.81). The DeLong test indicated that the combined model significantly improved discrimination of Aβ positivity (*p* = .039).

### Sensitivity analyses

#### Stepwise biomarker selection with age adjustment

The same plasma biomarkers identified in the primary analysis were retained after including age in the stepwise selection procedure. Specifically, BD-p-tau217 (B = 0.22; 95% CI, 0.19 to 0.25; *p* < .001), Aβ42 (B = −0.08; 95% CI, −0.11 to −0.06; *p* < .001), IGFBP7 (B = −0.04; 95% CI, −0.08 to −0.01; *p* = .006) and BACE1 (B = −0.08; 95% CI, −0.08 to −0.01; *p* < .001) remained associated with global Aβ SUVR.

#### LASSO-selected plasma biomarkers associated with brain Aβ burden

The final LASSO model for plasma biomarker selection was evaluated at λ_min_ = 0.0122, corresponding to the step that minimized the cross-validated average square error. The model yielded results similar to those obtained using the stepwise approach, selecting BD-p-tau217 (B = 0.19), Aβ42 (B = −0.07), BACE1 (B = −0.03), and IGFBP7 (B = −0.02) as plasma biomarkers associated with global Aβ SUVR. Notably, plasma CD63 exhibited an unstable selection pattern across cross-validation iterations and was excluded from the final LASSO model at λ_min_ to minimize the cross-validated mean squared error.

## Discussion

In a community-based sample of Hispanic, non-Hispanic Black and non-Hispanic White individuals, we found that (i) BD-p-tau217 showed the strongest association with brain Aβ deposition and Aβ positivity among the evaluated plasma biomarkers; (ii) the association between BD-p-tau217 and brain Aβ deposition was attenuated in individuals with moderately/severely decreased kidney function or failure; (iii) lower plasma Aβ42, BACE1, and IGFBP7 levels were independently associated with greater brain Aβ levels and combining these biomarkers with BD-p-tau217 further improved the characterization of brain Aβ deposition; and (iv) incorporating BD-p-tau217 with Aβ42 and BACE1 was also associated with improved differentiation of Aβ PET positivity.

Previous studies demonstrated that NULISAseq-derived plasma biomarkers, particularly BD-p-tau species,^32,33^ can accurately identify AD-related pathological changes in individuals at preclinical or symptomatic stages of AD.^42–44^ Extending these findings into a community-based sample of CU individuals, we found significant correlations between several NULISAseq biomarkers and brain Aβ pathology. Specifically, higher levels of plasma BD-p-tau217, BD-p-tau181, and total p-tau217, as well as lower levels of plasma Aβ42 and IGFBP7, were correlated with greater brain Aβ deposition. Moreover, these biomarkers were inversely correlated with eGFR; yet their correlations with brain Aβ deposition remained independent of eGFR. While similar findings have been reported previously, most studies have evaluated these relationships using creatinine-based eGFR as a measure of kidney function.^27,26,45^ In contrast, we used cystatin C-based eGFR, which is less affected by body weight, muscle mass, and diet compared with creatinine-based eGFR.^46,47^

BD-p-tau217 emerged as the plasma biomarker most strongly associated with brain Aβ deposition and Aβ PET positivity among conventional and novel NULISAseq biomarkers of AD pathology. Plasma Aβ42, a well-established biomarker of the Aβ proteinopathy pathway,^4^ demonstrated the second strongest association with brain Aβ deposition. Similarly, previous research found that BD-p-tau217 was more strongly associated with abnormal brain Aβ levels and exhibited superior classification performance for Aβ PET positivity compared with other plasma biomarkers of Aβ pathology, including total p-tau217,^32,33^ BD-p-tau181 and BD-p-tau231.^32^ While other brain-derived and total p-tau isoforms were excluded from our stepwise models to minimize multicollinearity, Spearman correlation analyses showed that BD-p-tau217 had stronger correlations with brain Aβ burden compared with total p-tau217 and BD-p-tau181, further supporting the prior evidence.^32,33^ Notably, most existing studies have shown that BD-p-tau species can detect Aβ pathology in clinical settings.^28,32,33^ Our findings in a community-based sample further highlight the potential of plasma BD-p-tau217 to reflect early brain Aβ deposition at the population level.

Several studies reported that chronic conditions, such as CKD, may influence plasma biomarker levels as risk factors for AD or through physiological processes.^18,24,25^ In our study, levels of most plasma biomarkers increased significantly with advancing CKD stage, while brain Aβ burden or Aβ PET positivity did not differ significantly by CKD stage. These findings suggest that the observed elevations in plasma biomarkers may be driven by CKD-related physiological alterations, such as reduced protein clearance.^18,25^ Furthermore, we found that the association of BD-p-tau217 with brain Aβ burden was modified by CKD stage. While the strength of this association was similar among individuals with normal/high and mildly decreased kidney function, it was attenuated in individuals with moderately/severely decreased kidney function or failure compared to those with normal/high kidney function. Similarly, a recent study reported that an advanced CKD stage, defined as eGFR <45, may confound the utility of p-tau217-based measures as markers of brain Aβ deposition.^45^ Leveraging a brain-derived assay of p-tau217, our findings further suggest that significant kidney dysfunction (i.e., eGFR < 60)^46^ may also modify the association between BD-p-tau217 and brain Aβ pathology. Nevertheless, the impact of CKD on plasma p-tau217 levels may diminish in the presence of a greater AD pathology burden than observed in our community-based cohort, as previously reported for both BD-p-tau217^33^ and total p-tau217.^27^ In addition, our sample included a relatively small number of individuals with moderately/severely decreased kidney function or failure, which may have reduced the precision of estimates.^26^ Indeed, when participants with any degree of kidney dysfunction were combined into a single group and compared with those who had normal/high kidney function, no significant group differences were observed on the strength of the association between BD-p-tau217 and brain Aβ burden. Collectively, these findings highlight the need for future studies with more balanced representation across CKD stages to further evaluate the potential influence of CKD on the association between BD-p-tau217 and brain Aβ pathology.

Recent evidence suggests that assays with multianalyte measurement capabilities may provide a more comprehensive characterization of peripheral biological processes and facilitate the identification of novel AD plasma biomarkers.^42–44^ Utilizing NULISAseq multiplex assay, our stepwise analysis identified two novel plasma biomarkers—BACE1 and IGFBP7—that were inversely associated with brain Aβ deposition. These findings remained robust when biomarker selection was performed after adjusting for age or using the LASSO method. While elevated levels of BACE1^48^ and IGFBP7^49^ have been previously reported in individuals with mild cognitive impairment or AD dementia, their relationship with Aβ pathology remain largely unexplored in CU populations. Notably, a previous study comprising predominantly of CU individuals did not find any association between these biomarkers and brain Aβ burden.^42^ However, the previous study included older individuals with greater pathological burden relative to our sample,^42^ which may partly explain the discrepant findings. In our study, combining BACE1, IGFBP7, and Aβ42 with plasma BD-p-tau217 was also related to an enhanced characterization of brain Aβ deposition, as indicated by a significant improvement in model fit relative to BD-p-tau217 alone. Similarly, incorporating plasma BD-p-tau217 with Aβ42 and BACE1 achieved better performance for detecting Aβ PET positivity compared with plasma BD-p-tau217 alone, raising the AUC from 0.68 to 0.75. Consistent with this observation, previous studies demonstrated that combining plasma biomarkers of AD, such as plasma p-tau217 and Aβ42/40 ratio, can improve the detection of Aβ pathology in CU individuals and streamline screening of individuals for primary prevention and anti-amyloid trials.^14,16^ Notably, the overall discriminative performance of plasma biomarkers for Aβ positivity was modest in our sample, which could be expected given that plasma biomarkers typically show lower accuracy for detecting Aβ positivity in CU populations compared with cognitively impaired populations.^50^ Additionally, the prevalence of Aβ PET positivity was low (16%), which could impact biomarker classification performance.^20^ Overall, future studies in independent cohorts are warranted to validate the utility of plasma BACE1 and IGFBP7 as potential indicators of Aβ pathology.

Our study has several limitations. First, the cross-sectional design precludes inference about the ability of plasma biomarkers to predict Aβ accumulation. Second, we did not evaluate the performance of our assay relative to other available assays for BD-p-tau quantification, such as Lilly’s p-tau217 electrochemiluminescence immunoassay.^28^ Future cross-platform comparisons would provide further insight into the robustness and utility of NULISAseq-derived BD-p-tau217 for detecting Aβ pathology. Third, the effect sizes for the associations between plasma biomarkers and Aβ PET burden were small. However, this may be expected given that the performance of blood-based biomarker immunoassays is typically lower in preclinical compared with clinical stages of AD.^50^ Lastly, other conditions that may affect plasma biomarker levels, such as adiposity and stroke,^18,25^ were not evaluated in this study.

The main strength of our study is the evaluation of multiple established and novel NULISAseq AD pathology biomarkers, including recently developed BD-p-tau isoforms, in relation to early brain Aβ deposition in a relatively large, community-based sample. Additionally, we evaluated the extent to which kidney function may modify the link between plasma biomarkers and brain Aβ deposition using cystatin C-derived eGFR, a more precise marker of kidney function compared with creatinine-based eGFR.^46,47^ Finally, the racial and ethnic diversity of the sample enhances the generalizability of our findings and supports the broader applicability of NULISAseq biomarkers across heterogeneous populations.

In conclusion, our study identified BD-p-tau217 as the plasma biomarker most strongly associated with brain Aβ deposition in a diverse, community-based sample with low pathological burden. Notably, the association between BD-p-tau217 and brain Aβ deposition was attenuated in the presence of moderately/severely decreased kidney function or failure. Moreover, our study identified BACE1 and IGFBP7 as novel plasma biomarker candidates of brain Aβ deposition, which require replication in independent cohorts. Future studies are warranted to validate the utility of plasma BD-p-tau217—alone and in combination with other plasma biomarkers—for detecting brain Aβ deposition at the population level.

## Acknowledgements

This work was supported by United States National Institutes of Health grants R01AG050440 and R01AG055299 (Dr Luchsinger), and RF1AG051556 (Drs Luchsinger, Brickman and Moreno). Partial support was also provided by grants K24AG045334 (Dr Luchsinger), P30AG059303 (Dr Brickman), ULT1TR001873 (Dr Reilly). Dr Teresi is supported by National Institute on Aging Claude D. Pepper Older Americans Independence Center (National Institute on Aging, 1P30AG028741, Morrison and Aldrich) and the Alzheimer’s Disease Resource Center for Minority Aging Research (National Institute on Aging, P30AG066462, Manly and Brickman). Dr Lao is in part supported by grant R00AG065506 from the National Institute on Aging. The views expressed in this manuscript are those of the authors and do not necessarily represent the views of the National Institute of Aging; the National Institutes of Health; or the U.S. Department of Health and Human Services.

