## Supplemental Table 1 for "Association of total and brain-derived Alzheimer’s disease plasma biomarkers with brain amyloid deposition in a community-based sample"

**Supplementary Table 1** Linear regression results for interaction between plasma BD-p-tau217 and CKD stage (normal/high *vs.* any degree of kidney dysfunction) on global Aβ SUVR

| **Outcome:** Global Aβ SUVR | | |
| --- | --- | --- |
| **Independent Variable** | **B (95% CI)** | ***p* value** |
| BD-p-tau217*CKD stage (mildly/moderately/severely decreased kidney function or failure) | −0.06 (−0.14 to 0.020) | .139 |
| CKD stage (mildly/moderately/severely decreased kidney function or failure) | 0.72 (−0.21 to 1.65) | .129 |
| BD-p-tau217 | 0.27 (0.19 to 0.35) | <.001 |
| Aβ42 | −0.08 (−0.11 to −0.06) | <.001 |
| IGFBP7 | −0.04 (−0.07 to −0.009) | .002 |
| BACE1 | −0.05 (−0.08 to −0.02) | .012 |

Abbreviations: Aβ, amyloid-beta; SUVR, standardized uptake value ratio; BD-p-tau, brain-derived p-tau; IGFBP7, insulin-like growth-factor binding protein 7; BACE1, beta-secretase 1.
