## Supplemental Table 2 for "Association of total and brain-derived Alzheimer’s disease plasma biomarkers with brain amyloid deposition in a community-based sample"

**Supplementary Table 2** Stepwise selection of NULISAseq plasma biomarkers associated with Aβ PET positivity

| **Outcome:** Aβ PET positivity | | |  |
| --- | --- | --- | --- |
| **Plasma Biomarker** | **Odds Ratio (95% CI)** | ***p* value** | **Nagelkerke *R^2^*** |
| BD-p-tau217 | 23.7 (9.16 to 61.4) | <.001 | .101 |
| Aβ42 | 0.19 (0.10 to 0.35) | <.001 | .188 |
| BACE1 | 0.26 (0.10 to 0.64) | .003 | .213 |

Plasma biomarkers selected by stepwise logistic regression are listed in the order of selection. Nagelkerke *R*² values represent the performance of the cumulative model at each step. Abbreviations: Aβ, amyloid-beta; SUVR, standardized uptake value ratio; BD-p-tau, brain-derived p-tau; BACE1, beta-secretase.
